# Early effectiveness of the BNT162b2 KP.2 vaccine against COVID-19 in the US Veterans Affairs Healthcare System

**DOI:** 10.1101/2024.12.26.24319566

**Authors:** Haley J. Appaneal, Vrishali V. Lopes, Laura Puzniak, Evan J. Zasowski, Luis Jodar, John M. McLaughlin, Aisling R. Caffrey

## Abstract

We conducted a test-negative case-control study within the US Veterans Affairs Healthcare System to estimate early vaccine effectiveness (VE) of the BNT162b2 KP.2 vaccine (Pfizer 2024-2025 formulation) against COVID-19 outcomes. Among 44,598 acute respiratory infections between September 5 and November 30, 2024, VE was 68% (42-82%), 57% (46-65%), and 56% (36-69%) against COVID-19-associated hospitalizations, emergency department and urgent care visits, and outpatient visits, respectively. Uptake of updated COVID-19 vaccines remains low.

## BACKGROUND

On August 22, 2024, the US Food and Drug Administration (FDA) authorized and approved the use of the Omicron KP.2-adapted COVID-19 vaccine formulation for the 2024-2025 respiratory virus season. The US Centers for Disease Control and Prevention (CDC) recommends everyone ages 6 months and older receive an updated COVID-19 vaccine, regardless of previous COVID-19 vaccination history. Over the past several years, COVID-19 vaccines have been key in blunting the public health impact of SARS-CoV-2.^1–4^ However, COVID-19 vaccination rates have fallen over the past several years.^5^ Data demonstrating real-world effectiveness of KP.2-adapted COVID-19 vaccines during the early part of the 2024-2025 respiratory virus season may help improve vaccine acceptance for the remainder of this season’s COVID-19 vaccine campaign, as well as future campaigns. We evaluated early vaccine effectiveness (VE) of the BNT162b2 KP.2 vaccine (Pfizer 2024-2025 formulation) against COVID-19 hospitalizations, emergency department (ED) and urgent care (UC) visits, and outpatient visits.

## METHODS

### Ethics Approval

Our study complies with all relevant ethical regulations and was determined to be exempt by the VA Providence Healthcare System (VAPHS) Institutional Review Board (IRB) and approved by the VAPHS Research and Development Committee. As this was a retrospective study of existing health records and exempt from IRB review, informed consent requirements are not applicable.

### Setting and Participants

We conducted a nationwide test-negative case-control study using clinical data from patients of the US Veterans Affairs (VA) Healthcare System, the largest integrated healthcare system in the US. We assessed VE of BNT162b2 KP.2 vaccine among adult patients (≥18 years of age) diagnosed with an acute respiratory infection (ARI, **Supplemental Table 1**) in the hospital, ED/UC, or outpatient setting (in-person or virtual) between September 5, 2024 and November 30, 2024. As in our prior work,^6^ to be included patients had to be tested for SARS-CoV-2 via nucleic acid amplification test (NAAT) or rapid antigen test (RAT) within 14 days prior through 3 days after the ARI encounter and patients were excluded if they (1) did not have at least one visit to the VA Healthcare System in the previous 12 months, (2) had another prior positive SARS-CoV-2 test in the 90 days prior to their ARI episode, (3) received a KP.2 vaccine other than BNT162b2, (4) received BNT162b2 KP.2 vaccine within 8 weeks of a prior COVID-19 vaccine dose, (5) received BNT162b2 KP.2 vaccine within 14 days prior to their ARI episode, (6) received BNT162b2 KP.2 vaccine but the date of administration was unknown, or (7) received a COVID-19 antiviral (nirmatrelvir/ritonavir, remdesivir, or molnupiravir) within 30 days prior to their ARI episode. Patients could contribute more than one ARI episode to the study if the episodes were more than 30 days apart (**Supplemental Figure 1**).

### Outcomes

Within each ARI outcome category (hospitalizations, ED/UC visits, outpatient visits), cases were those with a positive SARS-CoV-2 test result, and controls were those who tested negative.

### Exposure

Exposed patients received the BNT162b2 KP.2 vaccine at least 14 days before the ARI encounter. Unexposed patients did not receive a KP.2 strain-adapted COVID-19 vaccine of any kind, regardless of prior COVID-19 vaccination history. Vaccine exposure status was determined from the VA’s integrated electronic health record, which captures vaccines administered both within and outside of the VA Healthcare System.^7^ COVID-19 vaccines were offered free of charge to all Veterans enrolled in the VA Healthcare System based on CDC recommendations at the time of study conduct.^8^

### Statistical Analyses

Separate multivariable logistic regression models were used to compare the odds of receiving BNT162b2 KP.2 vaccine among SARS-CoV-2 positive cases and test-negative controls within each ARI outcome category, while adjusting for potentially confounding variables. We selected the following variables to control for a priori based on previous literature and previous work: age (18–64, 65–74, or ≥75 years), sex (male or female), race (Black, White, or other race), ethnicity (Hispanic or non-Hispanic), body mass index (underweight, healthy weight, overweight, obese, or missing), Charlson Comorbidity Index (0, 1, 2, 3, or ≥4), smoking status (current/former smoker or never smoker/unknown), immunocompromised status (yes or no), receipt of pneumococcal vaccine in the past 5 years (yes or no), number of interactions with the VA healthcare system in the year prior (hospital admission, nursing home admission, ED/UC visit, primary care visit; 0 or ≥1 for each), evidence of documented prior SARS-CoV-2 infection (yes or no), and US Census region (Northeast, Midwest, South, or West). We checked for assumptions and model fit in all logistic regression models. To calculate VE, we subtracted the corresponding adjusted odds ratios (OR) and 95% confidence intervals (CI) from 1 and multiplied by 100%. Subgroup analyses were conducted in those ≥65 years of age. All analyses were conducted using SAS (Version 9.4 and Enterprise Guide 8.3, SAS Institute Inc., Cary, NC, USA).

## DATA AVAILABILITY

The data supporting the findings of this study are not publicly available due to the inclusion of identifiable protected health information from the Veterans Health Administration. Privacy regulations prevent the open sharing of the individual-level data used in this study and any data covered under these regulations cannot be shared. The Veterans Health Administration may approve the sharing of some study data after verifying de-identification, though this may not include all final study data. Each request is subject to approval by the ethics board, privacy office, and information systems and security office. For such requests, please contact the corresponding author.

## RESULTS

This study included 44,598 ARI encounters with valid SARS-CoV-2 test results (**Supplemental Figure 1**), of which 9,439 (21.2%) were hospital admissions, 23,966 (53.7%) were ED/UC visits, and 11,193 (25.1%) were outpatient visits. Median age was 68 years (58.6% were ≥65 years of age), 87.7% were male, and 73.9% had at least one chronic medical condition. Overall, 16.2% (7,224) tested positive for SARS-CoV-2, and 1,666 (3.7%) received the BNT162b2 KP.2 vaccine, with a median time since receipt of 33 days (interquartile range 22-46). Overall, 138 of 7224 cases (1.9%) and 1,528 of 37,374 controls (4.1%) received the BNT162b2 KP.2 vaccine (Table 1).

**Table 1.**
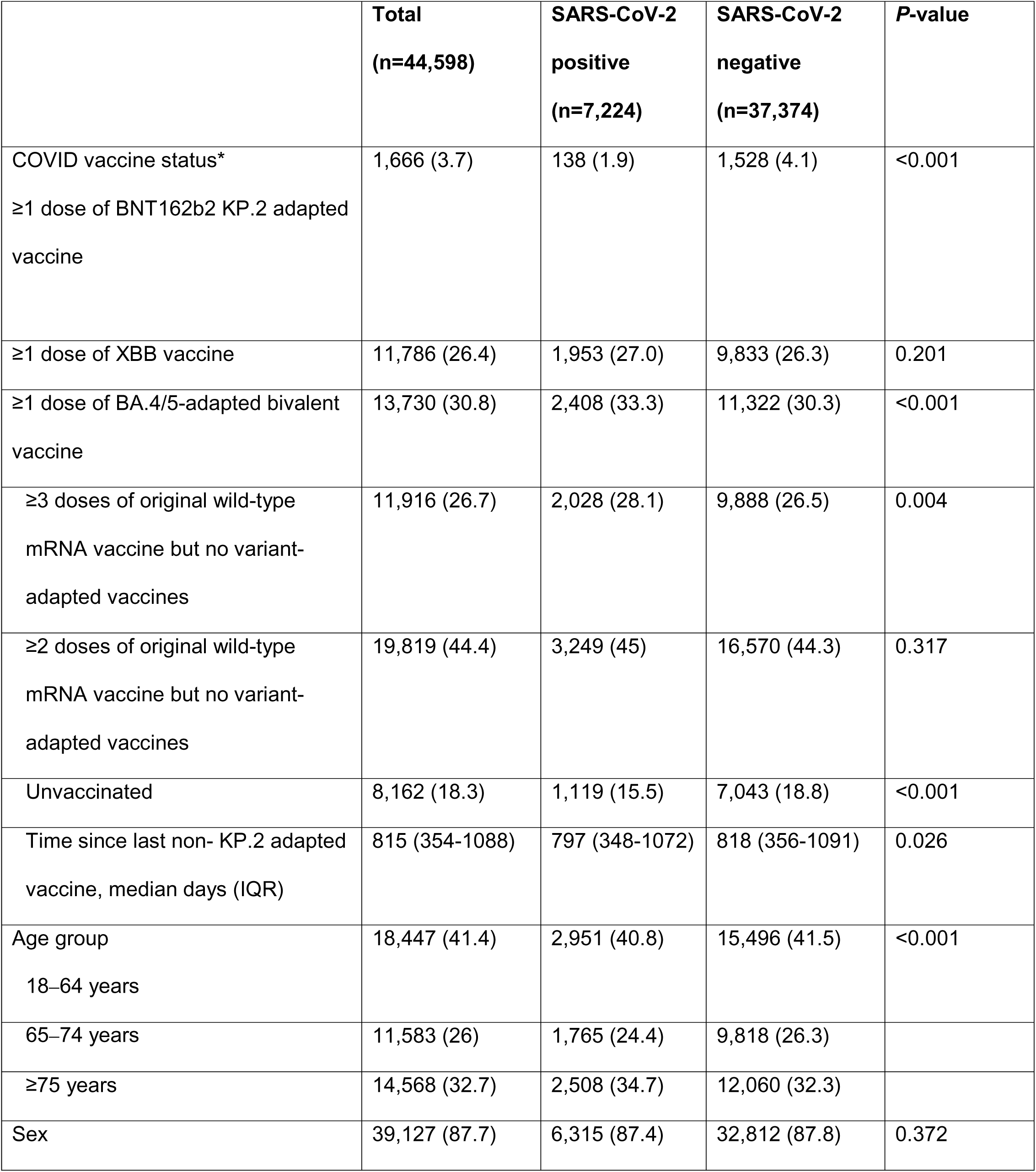
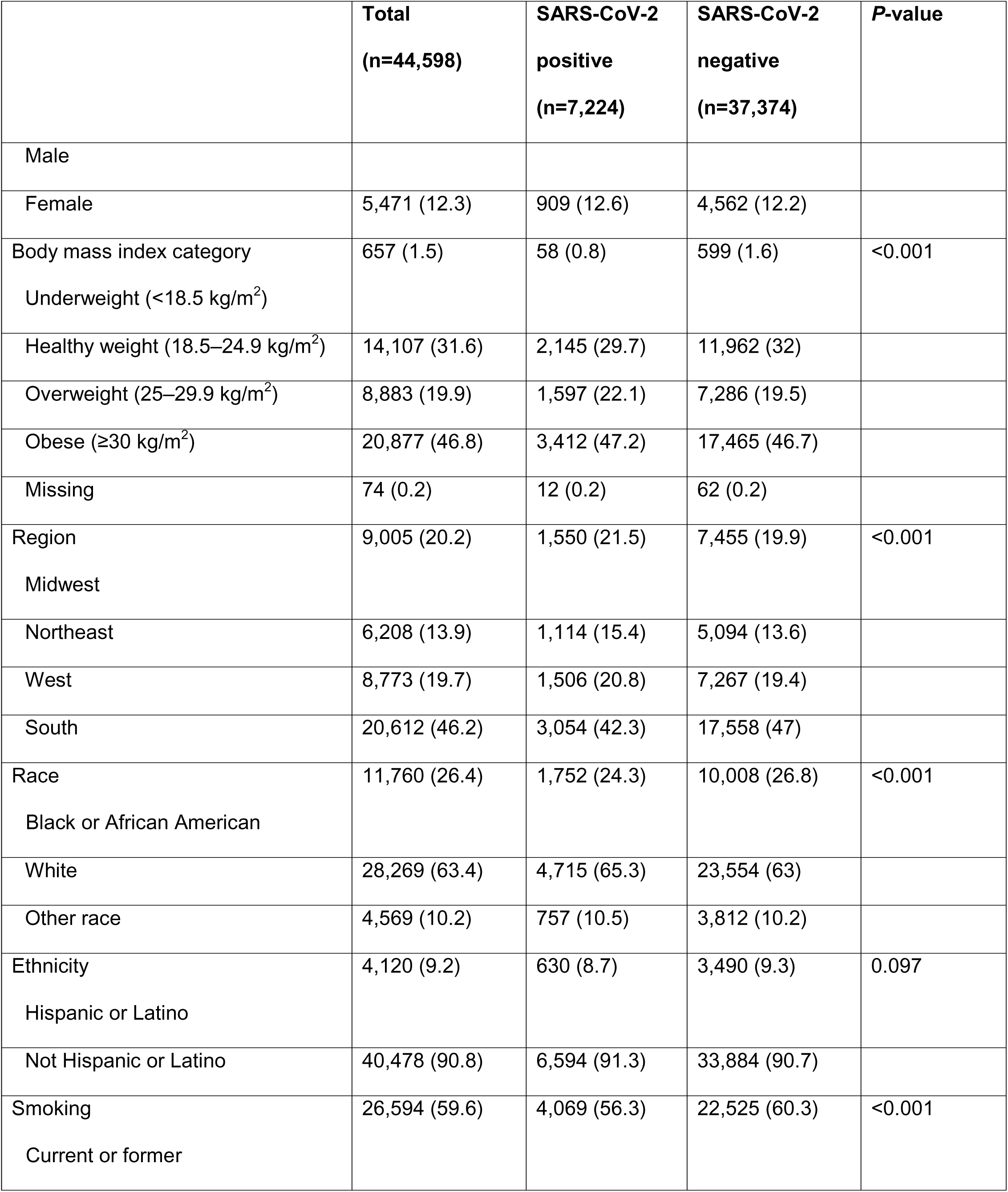
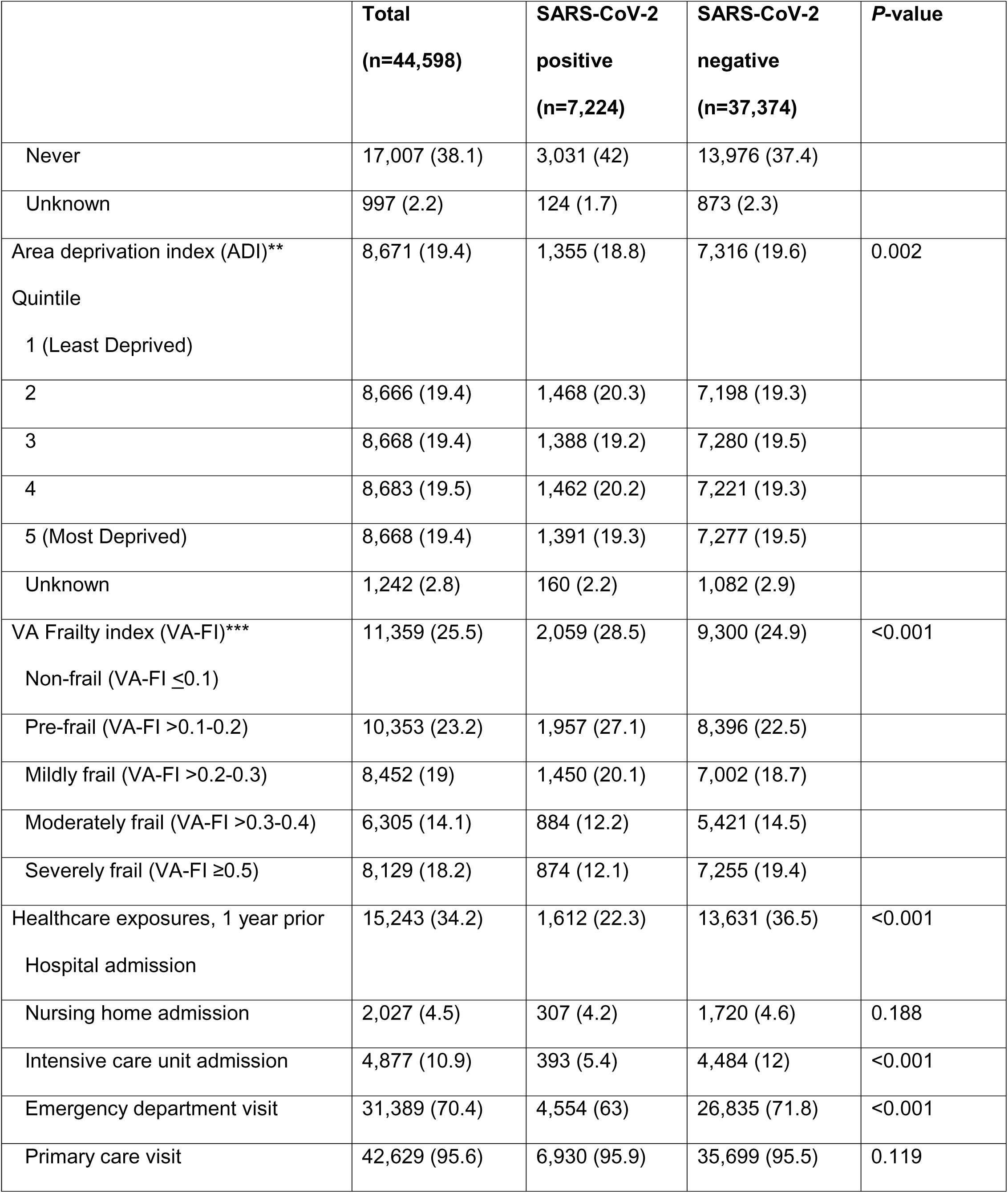
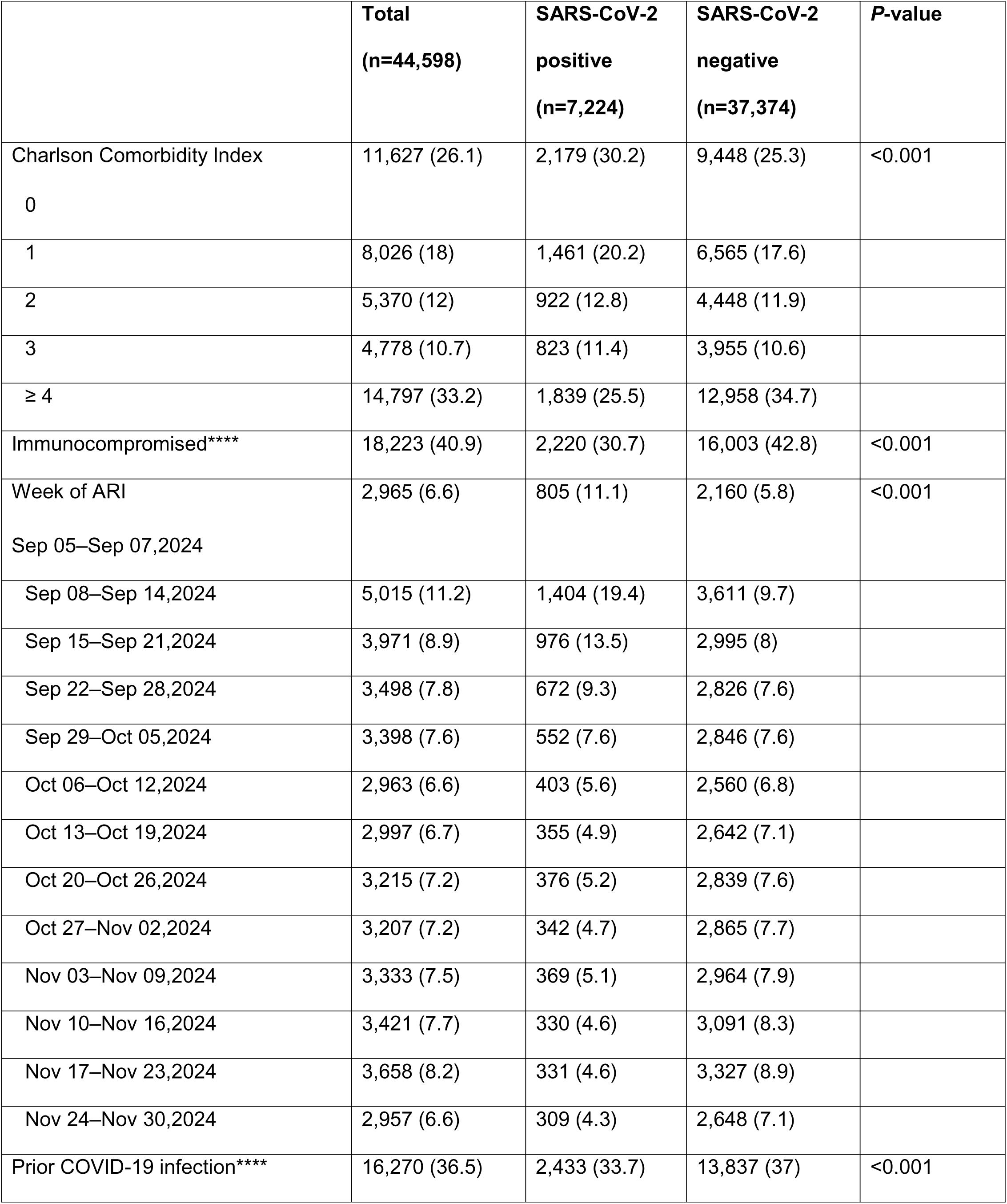
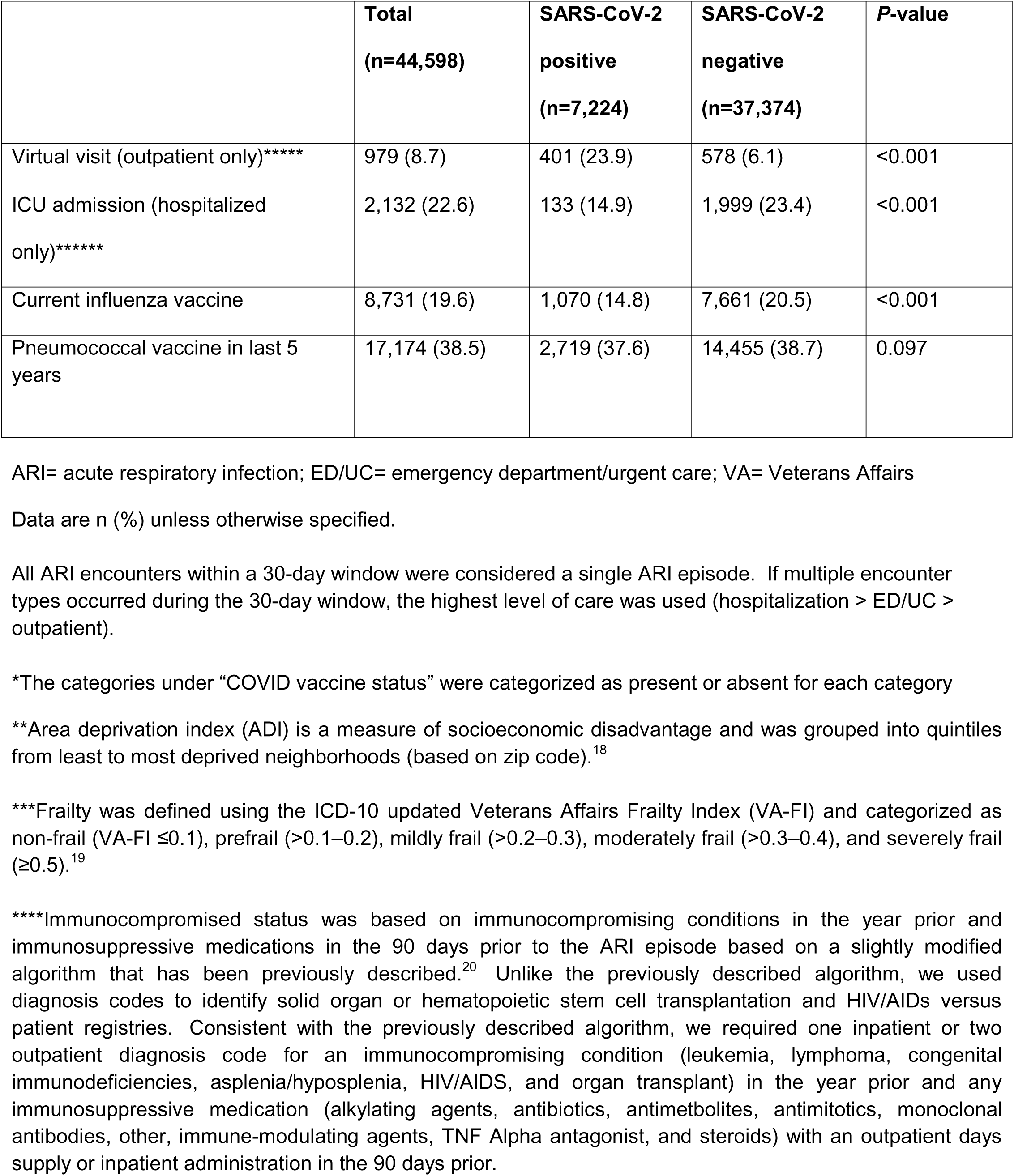
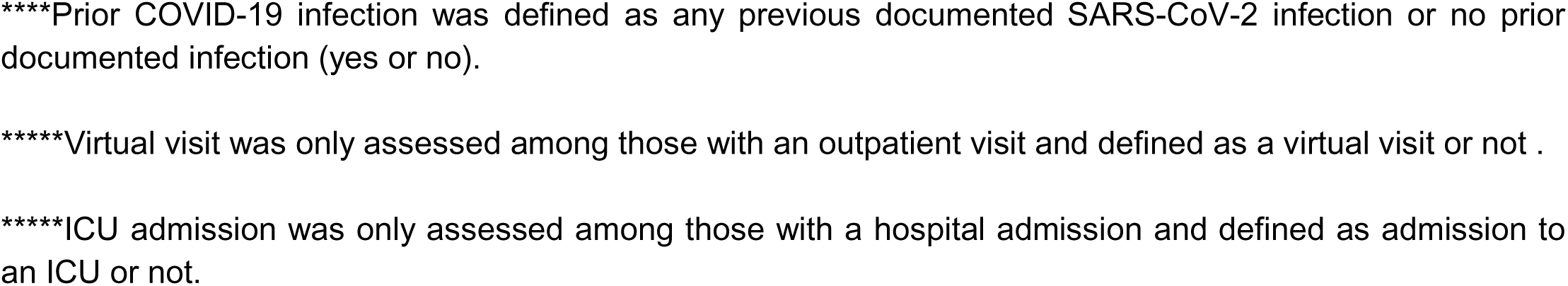
Demographics and clinical characteristics of acute respiratory infection encounters by case-control status.

Compared to those who tested positive for SARS-CoV-2, test-negative controls had a higher proportion of individuals with a Charlson Comorbidity Index ≥4 (34.7% *vs* 25.5%; *P*<.001), hospital admission in the prior year (36.5% *vs* 22.3%; *P*<.001), an ED visit in the prior year (71.8% *vs* 63.0%; *P*<.001), and prior documented SARS-CoV-2 infection (37.0% *vs* 33.7%; *P*<.001). These and other differences by case-control status are described in **Table 1** and **Supplemental Table 2**. Demographics and clinical characteristics by case-control status for each VE outcome are presented in **Supplemental Tables 3-5** and by BNT162b2 KP.2 vaccination status in **Supplemental Table 6**.

Overall, adjusted VE of the BNT162b2 KP.2 vaccine (compared to not receiving a KP.2 strain-adapted vaccine of any kind) against all COVID-19 outcomes was 56% (95% CI: 48–63%). Adjusted VE estimates by outcome were 68% (42–82%), 57% (46–65%), and 56% (36–69%) against hospitalizations, ED/UC visits, and outpatient visits, respectively (**Figure 1**). Among those ≥65 years of age, adjusted VE estimates by outcome were 75% (49–88%), 56% (44–66%), and 58% (36–73%) against hospitalizations, ED/UC visits, and outpatient visits, respectively (**Supplemental Table 7**).

**Figure 1.**
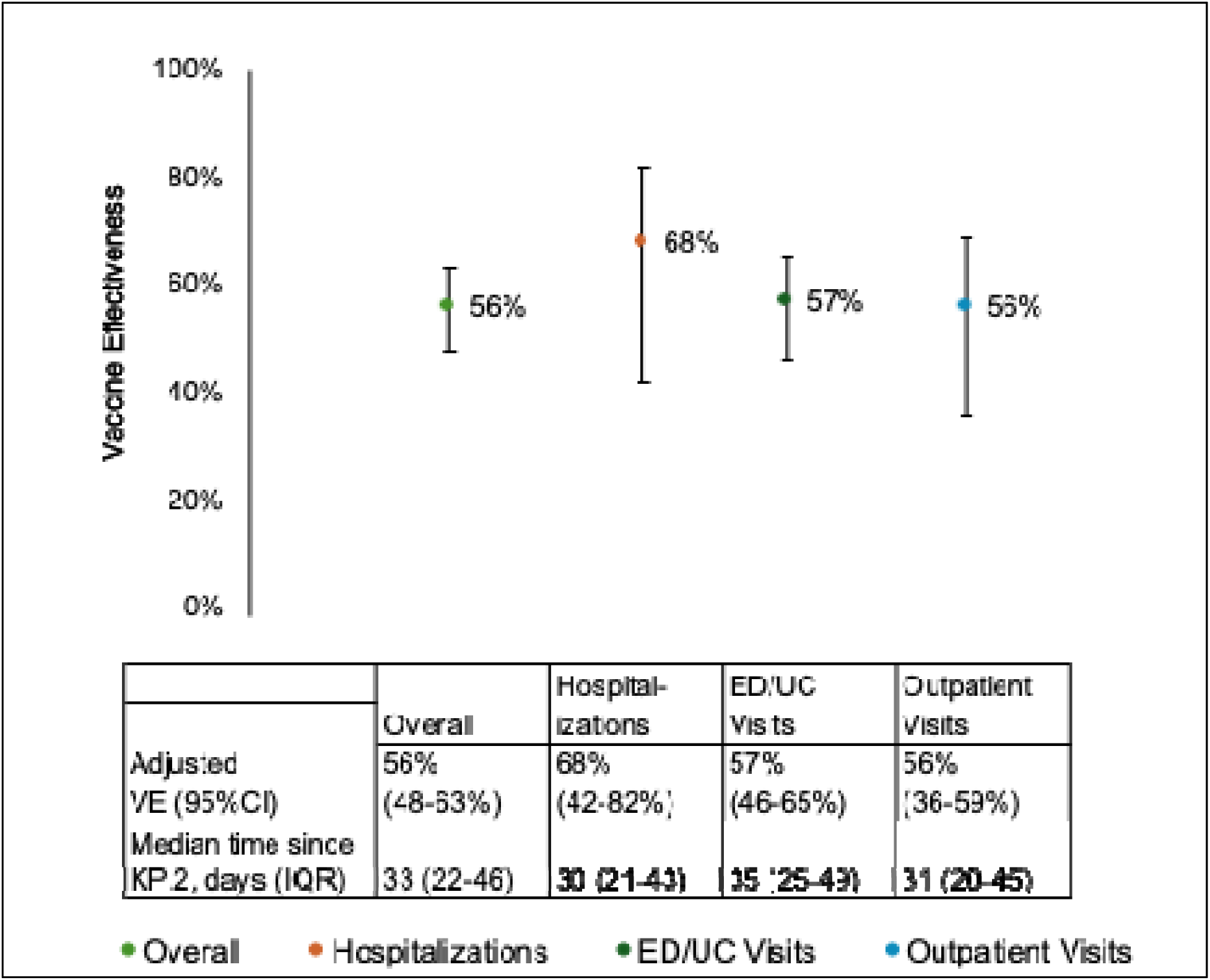
Adjusted effectiveness of the BNT162b2 KP.2 vaccine by COVID-19 outcomes ED/UC = emergency department/urgent care; IQR = interquartile range; KP.2 = BNT162b2 KP.2 adapted vaccine; VE= vaccine effectiveness Compared the odds of receiving the 2024/2025 BNT162b2 KP.2 strain-adapted COVID-19 vaccine between SARS-CoV-2 positive cases and SARS-CoV-2 negative controls. Adjusted for age (18–64, 65–74, ≥75 years), sex (male or female), race (Black, White, or other race), ethnicity (Hispanic or non-Hispanic), body mass index (BMI) categories (underweight, healthy weight, overweight, obese, missing), Charlson Comorbidity Index (0, 1, 2, 3, ≥4), receipt of pneumococcal vaccine in the past 5 years (yes or no), hospital admission, nursing home admission, ED/UC visit, primary care visit; 0 or ≥1 for each), prior documented SARS-CoV-2 infection (yes or no), smoking status (current/former smoker or never smoker/unknown), immunocompromised (yes or no), and Census region (Northeast, Midwest, South, or West).

## DISCUSSION

Our study is among the first to present early COVID-19 VE data for the 2024-2025 viral respiratory season. Results suggest that the BNT162b2 KP.2 vaccine provided significant protection against both mild and severe COVID-19 outcomes during the early part of the season. VE against milder outpatient disease (ED/UC or outpatient visits) was 561157% and against hospitalization was 68%. VE was similar when analyses were restricted to those ≥65 years of age, suggesting the vaccine provided comparable protection against the range of COVID-19 outcomes we studied in older adults who remain at increased risk of severe COVID-19. These current estimates are similar to our estimates of BNT162b2 XBB VE last year against the same outcomes and in the same population during a similar time period (September 25 to November 30, 2023).^6^ These findings suggest that early protection of annual COVID-19 vaccination over the last two years has been similar (VE of roughly 501170%, with the highest levels of protection seen against more severe outcomes like hospitalization).

According to CDC, however, uptake of KP.2 vaccines during the 2024-2025 respiratory virus season remains relatively low. As of December 12, 2024, only 21% of all adults ≥18 years of age, and 45% of adults ≥65 years reported receiving an updated KP.2 vaccine.^9^ These percentages are notably lower than influenza vaccination rates during a similar time period, with 41% and 67% of adults ≥18 and ≥65 years of age receiving the flu vaccine. Public health efforts should focus on improving acceptance and uptake of COVID-19 vaccines, as a recent report suggested that nearly 70% of US adults currently express hesitation or unwillingness toward receiving an updated COVID-19 vaccine.^9^

Similar to our previous work, this study has several limitations.^10^ In brief, the test-negative case-control study is considered a reliable design for evaluating real-world VE, but it is still susceptible to selection bias.^11–13^ Although we adjusted for key patient clinical and sociodemographic characteristics, there still may be residual confounding by unknown or unmeasured factors. We presented early VE estimates with a median time since KP.2 vaccination of only 33 days, thus future studies to evaluate longer-term durability are needed. Additionally, some ARI episodes may have involved seeking care "with COVID-19" rather than "for COVID-19," potentially leading to an underestimation of VE. Further, we were unable to conduct stratified analyses by specific vaccination history due to extensive heterogeneity in the number and type of previous COVID-19 vaccinations received. Previous studies, however, have suggested that older versions of COVID-19 vaccines likely offer little additional protection during subsequent seasons.^14^ Finally, our results may not be broadly generalizable, as they were conducted among the VA population which is generally older, predominantly male, and with a higher prevalence of multiple comorbid conditions compared to the general US or other global populations.^6,15^

In summary, the BNT162b2 KP.2 vaccine was 561168% effective at preventing a range of COVID-19 outcomes during the early part of the 2024-2025 respiratory virus season. Maintaining protection against COVID-19 with vaccination continues to be critical given that the post-pandemic burden of COVID-19 remains substantial. For example, cumulative rates of COVID-19-related hospitalization from September 2023 through October 2024 were estimated to be 242 and 821 per 100,000 among US adults ≥18 and ≥65 years of age,^16^ respectively, which were higher than hospitalization rates for influenza and respiratory syncytial virus during the same period.^17^ Despite this persistent burden and the increased protection observed with receiving updated vaccines over the last two respiratory virus seasons, uptake of updated COVID-19 vaccines remains low, even among older adults. Additional efforts to improve COVID-19 vaccine uptake to match that of annual influenza vaccine coverage are needed.

## ACKNOWLEDGEMENTS

**a. Funding**

This study was conducted as a collaboration between the University of Rhode Island (URI), VA Providence Healthcare System, and Pfizer. Pfizer is the study sponsor. URI and VA Providence Healthcare System received funding from Pfizer in connection with the development of this manuscript and for data analysis.

**b. Other Acknowledgements**

The views expressed are those of the authors and do not necessarily reflect the position or policy of the United States Department of Veterans Affairs.

The research study would not have been possible without the health information from patients under the care of the Veterans Health Administration. We express our gratitude to the VA patients for their invaluable contributions to medical and scientific progress.

## Author Contributions Statement

Conception and design of the study: Haley J. Appaneal, Vrishali V. Lopes, Laura Puzniak, Evan J. Zasowski, John M. McLaughlin, Aisling R. Caffrey

Data generation: Haley J. Appaneal, Vrishali V. Lopes, Aisling R. Caffrey

Analysis and/or interpretation of the data: Haley J. Appaneal, Vrishali V. Lopes, Laura Puzniak, Evan J. Zasowski, Luis Jodar, John M. McLaughlin, Aisling R. Caffrey

Preparation or critical revision of the manuscript: Haley J. Appaneal, Vrishali V. Lopes, Laura Puzniak, Evan

J. Zasowski, Luis Jodar, John M. McLaughlin, Aisling R. Caffrey

## Competing Interests Statement

Haley J. Appaneal has received research funding from Pfizer.

Vrishali V. Lopes has no competing interest to declare.

Laura Puzniak, Evan J. Zasowski, Luis Jodar, and John M. McLaughlin are employees and shareholders of Pfizer Inc.

Aisling R. Caffrey has received research funding from AbbVie, Merck, and Pfizer.

