## Supplemental Materials for "Early effectiveness of the BNT162b2 KP.2 vaccine against COVID-19 in the US Veterans Affairs Healthcare System"

**Supplemental Figure 1.** Study selection criteria

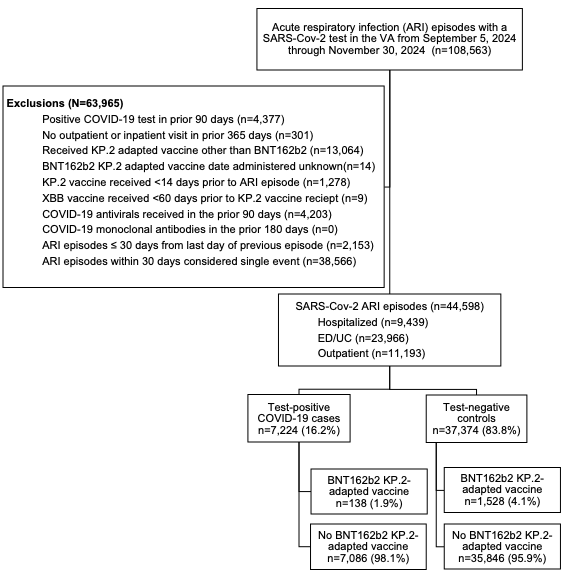

ED/UC= emergency department/urgent care; VA = Veterans Affairs

Patients could contribute more than one ARI episode to the study if the episodes were more than 30 days apart.

**Supplemental Table 1.** Acute respiratory infection diagnosis codes (ICD-10)

| **ICD-10 Code** | **Diagnosis** |
| --- | --- |
| A22.1 | Pulmonary anthrax |
| A37.00 | Whooping cough due to *Bordetella pertussis* without pneumonia |
| A37.01 | Whooping cough due to *Bordetella pertussis* with pneumonia |
| A37.10 | Whooping cough due to *Bordetella parapertussis* without pneumonia |
| A37.11 | Whooping cough due to *Bordetella parapertussis* with pneumonia |
| A37.80 | Whooping cough due to other Bordetella species without pneumonia |
| A37.81 | Whooping cough due to other Bordetella species with pneumonia |
| A37.90 | Whooping cough, unspecified species without pneumonia |
| A37.91 | Whooping cough, unspecified species with pneumonia |
| A48.1 | Legionnaires' disease |
| B25.0 | Cytomegaloviral pneumonitis |
| B34.2 | Coronavirus infection, unspecified |
| B44.0 | Invasive pulmonary aspergillosis |
| B77.81 | Ascariasis pneumonia |
| B97.29 | Other coronavirus as the cause of diseases classified elsewhere |
| J00* | Acute nasopharyngitis [common cold] |
| J01* | Acute sinusitis |
| J02* | Acute pharyngitis |
| J03* | Acute tonsillitis |
| J04* | Acute laryngitis and tracheitis |
| J05* | Acute obstructive laryngitis [croup] and epiglottitis |
| J06* | Acute upper respiratory infections of multiple and unspecified sites |
| J09.X1 | Influenza due to novel influenza a virus with pneumonia |
| J09.X2 | Influenza due to identified novel influenza A virus with other respiratory manifestations |
| J09.X3 | Influenza due to identified novel influenza A virus with gastrointestinal manifestations |
| J09.X9 | Influenza due to identified novel influenza A virus with other manifestations |
| J10.00 | Influenza due to other identified influenza virus with pneumonia |
| J10.01 | Influenza due to other identified influenza virus with the same other identified influenza virus pneumonia |
| J10.08 | Influenza due to other identified influenza virus with other specified pneumonia |
| J10.1 | Influenza due to other identified influenza virus with other respiratory manifestations |
| J10.2 | Influenza due to other identified influenza virus with gastrointestinal manifestations |
| J10.8* | Influenza due to other identified influenza virus with other manifestations |
| J10.81 | Influenza due to other identified influenza virus with encephalopathy |
| J10.82 | Influenza due to other identified influenza virus with myocarditis |
| J10.83 | Influenza due to other identified influenza virus with otitis media |
| J10.89 | Influenza due to other identified influenza virus with other manifestations |
| J11.00 | Influenza due to unidentified influenza virus with pneumonia |
| J11.08 | Influenza due to unidentified influenza virus with specified pneumonia |
| J11.1 | Influenza due to unidentified influenza virus with other respiratory manifestations |
| J11.2 | Influenza due to unidentified influenza virus with gastrointestinal manifestations |
| J11.81 | Influenza due to unidentified influenza virus with encephalopathy |
| J11.82 | Influenza due to unidentified influenza virus with myocarditis |
| J11.83 | Influenza due to unidentified influenza virus with otitis media |
| J11.89 | Influenza due to unidentified influenza virus with other manifestations |
| J12.0 | Adenoviral pneumonia |
| J12.1 | Respiratory syncytial virus pneumonia |
| J12.2 | Parainfluenza virus pneumonia |
| J12.3 | Human metapneumovirus pneumonia |
| J12.81 | Pneumonia due to SARS-associated coronavirus |
| J12.82 | Pneumonia due to coronavirus disease 2019 |
| J12.89 | Other viral pneumonia |
| J12.9 | Viral pneumonia, unspecified |
| J13 | Pneumonia due to *Streptococcus pneumoniae* |
| J14 | Pneumonia due to *Hemophilus influenzae* |
| J15.0 | Pneumonia due to *Klebsiella pneumoniae* |
| J15.1 | Pneumonia due to Pseudomonas |
| J15.20 | Pneumonia due to Staphylococcus, unspecified |
| J15.211 | Pneumonia due to methicillin susceptible *Staphylococcus aureus* |
| J15.212 | Pneumonia due to methicillin resistant *Staphylococcus aureus* |
| J.15.29 | Pneumonia due to other Staphylococcus |
| J15.3 | Pneumonia due to Streptococcus, group b |
| J15.4 | Pneumonia due to other Streptococci |
| J15.5 | Pneumonia due to *Escherichia coli* |
| J15.6 | Pneumonia due to other aerobic gram-negative bacteria |
| J15.7 | Pneumonia due to *Mycoplasma pneumoniae* |
| J15.8 | Pneumonia due to other specified bacteria |
| J15.9 | Unspecified bacterial pneumonia |
| J16.0 | Chlamydial pneumonia |
| J16.8 | Pneumonia due to other specified infectious organisms |
| J17 | Pneumonia in diseases classified elsewhere |
| J18.0 | Bronchopneumonia, unspecified organism |
| J18.1 | Lobar pneumonia, unspecified organism |
| J18.2 | Hypostatic pneumonia, unspecified organism |
| J18.8 | Other pneumonia, unspecified organism |
| J18.9 | Pneumonia, unspecified organism |
| J20.0 | Acute bronchitis due to *Mycoplasma pneumoniae* |
| J20.1 | Acute bronchitis due to *Hemophilus influenzae* |
| J20.2 | Acute bronchitis due to Streptococcus |
| J20.3 | Acute bronchitis due to coxsackievirus |
| J20.4 | Acute bronchitis due to parainfluenza virus |
| J20.5 | Acute bronchitis due to respiratory syncytial virus |
| J20.6 | Acute bronchitis due to rhinovirus |
| J20.7 | Acute bronchitis due to echovirus |
| J20.8 | Acute bronchitis due to other specified organisms |
| J20.9 | Acute bronchitis, unspecified |
| J21.* | Acute bronchiolitis |
| J21.0 | Acute bronchiolitis due to respiratory syncytial virus |
| J21.1 | Acute bronchiolitis due to human metapneumovirus |
| J21.8 | Acute bronchiolitis due to other specified organisms |
| J21.9 | Acute bronchiolitis, unspecified |
| J22 | Unspecified acute lower respiratory infection |
| J80 | Acute respiratory distress syndrome |
| J96.00 | Acute respiratory failure unspecified whether with hypoxia or hypercapnia |
| J96.01 | Acute respiratory failure with hypoxia |
| J96.02 | Acute respiratory failure with hypercapnia |
| J96.10 | Chronic respiratory failure, unspecified with hypoxia or hypercapnia |
| J96.11 | Chronic respiratory failure with hypoxia |
| J96.12 | Chronic respiratory failure with hypercapnia |
| J96.20 | Acute and chr resp failure, unspecified with hypoxia or hypercapnia |
| J96.21 | Acute and chronic respiratory failure with hypoxia |
| J96.22 | Acute and chronic respiratory failure with hypercapnia |
| J96.90 | Respiratory failure, unspecified, unspecified with hypoxia or hypercapnia |
| J96.91 | Respiratory failure, unspecified with hypoxia |
| J96.92 | Respiratory failure, unspecified with hypercapnia |
| M35.81 | Multisystem inflammatory syndrome |
| R04.2 | Hemoptysis |
| R05 | Cough |
| R05.1 | Acute cough |
| R05.2 | Subacute cough |
| R05.3 | Chronic cough |
| R05.4 | Cough syncope |
| R05.8 | Other specified cough |
| R05.9 | Cough, unspecified |
| R06.00 | Dyspnea/abnormalities of breathing unspecified |
| R06.02 | Shortness of breath |
| R06.03 | Acute respiratory distress |
| R06.09 | Other forms of dyspnea |
| R06.1 | Stridor |
| R06.82 | Tachypnea, not elsewhere classified |
| R06.89 | Other abnormalities of breathing |
| R07.1 | Chest pain on breathing |
| R09.0* | Asphyxia and hypoxemia |
| R09.01 | Asphyxia |
| R09.02 | Hypoxemia |
| R09.1 | Pleurisy |
| R09.2 | Respiratory arrest |
| R50.9 | Fever, unspecified |
| U04* | SARS (WHO 2019) |
| u04.9 | SARS, unspecified (WHO 2019) |
| U07.1 | COVID-19 |
| U07.2 | COVID-19, virus not identified (clinically diagnosed) |

**Supplemental Table 2.** Medical history of acute respiratory infection encounters by case-control status

|  | **Total (n=44,598)** | **SARS-CoV-2 positive**  **(n=7,224)** | **SARS-CoV-2 negative**  **(n=37,374)** | ***P*-value** |
| --- | --- | --- | --- | --- |
| Medical History  Acute cerebrovascular disease | 2,673 (6) | 322 (4.5) | 2,351 (6.3) | <0.001 |
| Acute myocardial infarction | 2,035 (4.6) | 164 (2.3) | 1,871 (5) | <0.001 |
| Alcohol and substance related disorders | 12,323 (27.6) | 1,616 (22.4) | 10,707 (28.6) | <0.001 |
| Any cancer or malignancy | 19,271 (43.2) | 2,984 (41.3) | 16,287 (43.6) | <0.001 |
| Aortic and peripheral arterial embolism or thrombosis | 246 (0.6) | 21 (0.3) | 225 (0.6) | 0.001 |
| Asthma | 4,590 (10.3) | 642 (8.9) | 3,948 (10.6) | <0.001 |
| Benign prostatic hyperplasia | 11,427 (25.6) | 1,724 (23.9) | 9,703 (26) | <0.001 |
| Cardiac dysrhythmias | 13,229 (29.7) | 1,754 (24.3) | 11,475 (30.7) | <0.001 |
| Chronic kidney disease | 6,642 (14.9) | 850 (11.8) | 5,792 (15.5) | <0.001 |
| Chronic obstructive pulmonary disease and bronchiectasis | 12,986 (29.1) | 1,473 (20.4) | 11,513 (30.8) | <0.001 |
| Congestive heart failure | 9,040 (20.3) | 932 (12.9) | 8,108 (21.7) | <0.001 |
| Coronary atherosclerosis and other heart disease | 11,881 (26.6) | 1,662 (23) | 10,219 (27.3) | <0.001 |
| Delirium, dementia, and other cognitive disorders | 5,170 (11.6) | 625 (8.7) | 4,545 (12.2) | <0.001 |
| Diabetes with or without chronic complications | 20,884 (46.8) | 3,346 (46.3) | 17,538 (46.9) | 0.343 |
| Epilepsy | 1,520 (3.4) | 187 (2.6) | 1,333 (3.6) | <0.001 |
| Human immunodeficiency virus (HIV) infection | 593 (1.3) | 74 (1) | 519 (1.4) | 0.013 |
| Hypertension | 28,801 (64.6) | 4,624 (64) | 24,177 (64.7) | 0.268 |
| Influenza | 1,245 (2.8) | 185 (2.6) | 1,060 (2.8) | 0.194 |
| Liver diseases | 5,123 (11.5) | 726 (10) | 4,397 (11.8) | <0.001 |
| Mental health conditions | 23,855 (53.5) | 3,610 (50) | 20,245 (54.2) | <0.001 |
| Osteoarthritis | 10,803 (24.2) | 1,805 (25) | 8,998 (24.1) | 0.098 |
| Peripheral and visceral atherosclerosis | 4,744 (10.6) | 585 (8.1) | 4,159 (11.1) | <0.001 |
| Pneumonia | 5,829 (13.1) | 517 (7.2) | 5,312 (14.2) | <0.001 |
| Pulmonary heart disease | 4,126 (9.3) | 396 (5.5) | 3,730 (10) | <0.001 |
| Rheumatoid arthritis | 1,063 (2.4) | 164 (2.3) | 899 (2.4) | 0.49 |
| Septicemia | 3,005 (6.7) | 251 (3.5) | 2,754 (7.4) | <0.001 |
| Thyroid disorder | 6,325 (14.2) | 986 (13.6) | 5,339 (14.3) | 0.156 |
| Tuberculosis | 126 (0.3) | 15 (0.2) | 111 (0.3) | 0.19 |

ARI= acute respiratory infection; ED/UC= emergency department/urgent care; VA= Veterans Affairs

Data are n (%) unless otherwise specified.

All ARI encounters within a 30-day window were considered a single ARI episode. If multiple encounter types occurred during the 30-day window, the highest level of care was used (hospitalization > ED/UC > outpatient).

Medical history included underlying conditions and diagnoses in the year prior to the ARI episode, identified using international classification of diseases (ICD)-10 codes.

**Supplemental Table 3.** Demographics and clinical characteristics of acute respiratory infection hospitalizations by case-control status

|  | **Total (n=9,439)** | **SARS-CoV-2 positive**  **(n=892)** | **SARS-CoV-2 negative**  **(n=8,547)** | ***P*-value** |
| --- | --- | --- | --- | --- |
| COVID vaccine status*  ≥1 dose of BNT162b2 KP.2 vaccine | 333 (3.5) | 12 (1.3) | 321 (3.8) | <0.001 |
| ≥1 dose of XBB vaccine | 2,909 (30.8) | 266 (29.8) | 2,643 (30.9) | 0.497 |
| ≥1 dose of BA.4/5-adapted bivalent vaccine | 3,321 (35.2) | 301 (33.7) | 3,020 (35.3) | 0.344 |
| ≥3 doses of original wild-type mRNA vaccine but no variant-adapted vaccines | 2,653 (28.1) | 276 (30.9) | 2,377 (27.8) | 0.048 |
| ≥2 doses of original wild-type mRNA vaccine but no variant-adapted vaccines | 4,082 (43.2) | 415 (46.5) | 3,667 (42.9) | 0.038 |
| Unvaccinated | 1,483 (15.7) | 121 (13.6) | 1,362 (15.9) | 0.064 |
| Time since last non-KP.2 adapted vaccine, median days (IQR) | 727 (337-1,058) | 751 (341-1,059) | 724 (337-1,057) | 0.375 |
| Age group  18–64 years | 1,850 (19.6) | 186 (20.9) | 1,664 (19.5) | 0.051 |
| 65–74 years | 2,997 (31.8) | 251 (28.1) | 2,746 (32.1) |  |
| ≥75 years | 4,592 (48.6) | 455 (51) | 4,137 (48.4) |  |
| Sex  Male | 8,897 (94.3) | 841 (94.3) | 8,056 (94.3) | 0.973 |
| Female | 542 (5.7) | 51 (5.7) | 491 (5.7) |  |
| Body mass index category  Underweight (<18.5 kg/m^2^) | 325 (3.4) | 22 (2.5) | 303 (3.5) | 0.114 |
| Healthy weight (18.5–24.9 kg/m^2^) | 4,068 (43.1) | 407 (45.6) | 3,661 (42.8) |  |
| Overweight (25–29.9 kg/m^2^) | 1,678 (17.8) | 169 (18.9) | 1,509 (17.7) |  |
| Obese (≥30 kg/m^2^) | 3,362 (35.6) | 293 (32.8) | 3,069 (35.9) |  |
| Missing | 6 (0.1) | <5 (<0.6) | 5 (0.1) |  |
| Region  Midwest | 1,807 (19.1) | 212 (23.8) | 1,595 (18.7) | <0.001 |
| Northeast | 1,268 (13.4) | 148 (16.6) | 1,120 (13.1) |  |
| West | 2,152 (22.8) | 186 (20.9) | 1,966 (23.0) |  |
| South | 4,212 (44.6) | 346 (38.8) | 3,866 (45.2) |  |
| Race  Black or African American | 2,236 (23.7) | 200 (22.4) | 2,036 (23.8) | 0.255 |
| White | 6,398 (67.8) | 604 (67.7) | 5,794 (67.8) |  |
| Other race | 805 (8.5) | 88 (9.9) | 717 (8.4) |  |
| Ethnicity  Hispanic or Latino | 721 (7.6) | 62 (7.0) | 659 (7.7) | 0.416 |
| Not Hispanic or Latino | 8,718 (92.4) | 830 (93.0) | 7,888 (92.3) |  |
| Smoking  Current or former | 6,426 (68.1) | 567 (63.6) | 5,859 (68.6) | <0.001 |
| Never smoked | 2,760 (29.2) | 313 (35.1) | 2,447 (28.6) |  |
| Unknown | 253 (2.7) | 12 (1.3) | 241 (2.8) |  |
| Area deprivation index (ADI)* Quintile  1 (Least Deprived) | 1,814 (19.2) | 192 (21.5) | 1,622 (19.0) | 0.599 |
| 2 | 1,699 (18) | 159 (17.8) | 1,540 (18.0) |  |
| 3 | 1,831 (19.4) | 164 (18.4) | 1,667 (19.5) |  |
| 4 | 1,856 (19.7) | 172 (19.3) | 1,684 (19.7) |  |
| 5 (Most Deprived) | 1,960 (20.8) | 178 (20.0) | 1,782 (20.8) |  |
| Missing | 279 (3.0) | 27 (3.0) | 252 (2.9) |  |
| VA Frailty index (VA-FI)**  Non-frail (VA-FI *≤*0.1) | 832 (8.8) | 81 (9.1) | 751 (8.8) | 0.014 |
| Pre-frail (VA-FI >0.1-0.2) | 1,484 (15.7) | 162 (18.2) | 1,322 (15.5) |  |
| Mildly frail (VA-FI >0.2-0.3) | 1,975 (20.9) | 207 (23.2) | 1,768 (20.7) |  |
| Moderately frail (VA-FI >0.3-0.4) | 1,978 (21.0) | 183 (20.5) | 1,795 (21.0) |  |
| Severely frail (VA-FI ≥0.5) | 3,170 (33.6) | 259 (29.0) | 2,911 (34.1) |  |
| Healthcare exposures, 1 year prior  Hospital admission | 5,428 (57.5) | 477 (53.5) | 4,951 (57.9) | 0.010 |
| Nursing home admission | 595 (6.3) | 65 (7.3) | 530 (6.2) | 0.204 |
| Intensive care unit admission | 1,873 (19.8) | 155 (17.4) | 1,718 (20.1) | 0.052 |
| Emergency department visit | 7,513 (79.6) | 685 (76.8) | 6,828 (79.9) | 0.029 |
| Primary care visit | 9,034 (95.7) | 851 (95.4) | 8,183 (95.7) | 0.636 |
| Charlson Comorbidity Index  0 | 808 (8.6) | 111 (12.4) | 697 (8.2) | <0.001 |
| 1 | 1,182 (12.5) | 111 (12.4) | 1,071 (12.5) |  |
| 2 | 1,137 (12.0) | 104 (11.7) | 1,033 (12.1) |  |
| 3 | 1,205 (12.8) | 126 (14.1) | 1,079 (12.6) |  |
| ≥4 | 5,107 (54.1) | 440 (49.3) | 4,667 (54.6) |  |
| Immunocompromised*** | 4,657 (49.3) | 332 (37.2) | 4,325 (50.6) | <0.001 |
| Week of ARI  Sep 05–Sep 07,2024 | 574 (6.1) | 75 (8.4) | 499 (5.8) | <0.001 |
| Sep 08–Sep 14,2024 | 1,036 (11.0) | 133 (14.9) | 903 (10.6) |  |
| Sep 15–Sep 21,2024 | 981 (10.4) | 116 (13.0) | 865 (10.1) |  |
| Sep 22–Sep 28,2024 | 886 (9.4) | 120 (13.5) | 766 (9.0) |  |
| Sep 29–Oct 05,2024 | 838 (8.9) | 81 (9.1) | 757 (8.9) |  |
| Oct 06–Oct 12,2024 | 723 (7.7) | 65 (7.3) | 658 (7.7) |  |
| Oct 13–Oct 19,2024 | 749 (7.9) | 59 (6.6) | 690 (8.1) |  |
| Oct 20–Oct 26,2024 | 758 (8) | 65 (7.3) | 693 (8.1) |  |
| Oct 27–Nov 02,2024 | 717 (7.6) | 49 (5.5) | 668 (7.8) |  |
| Nov 03–Nov 09,2024 | 721 (7.6) | 42 (4.7) | 679 (7.9) |  |
| Nov 10–Nov 16,2024 | 792 (8.4) | 51 (5.7) | 741 (8.7) |  |
| Nov 17–Nov 23,2024 | 565 (6) | 30 (3.4) | 535 (6.3) |  |
| Nov 24–Nov 30,2024 | 99 (1) | 6 (0.7) | 93 (1.1) |  |
| Prior COVID-19 infection**** | 3,179 (33.7) | 260 (29.1) | 2,919 (34.2) | 0.003 |
| ICU admission (hospitalized only)****** | 2,132 (22.6) | 133 (14.9) | 1,999 (23.4) | <0.001 |
| Current influenza vaccine | 1,980 (21.0) | 145 (16.3) | 1,835 (21.5) | <0.001 |
| Pneumococcal vaccine in last 5 years | 3,794 (40.2) | 339 (38.0) | 3,455 (40.4) | 0.161 |

ARI= acute respiratory infection; ED/UC= emergency department/urgent care; VA= Veterans Affairs

Data are n (%) unless otherwise specified.

All ARI encounters within a 30-day window were considered a single ARI episode. If multiple encounter types occurred during the 30-day window, the highest level of care was used (hospitalization > ED/UC > outpatient).

*The categories under “COVID vaccine status” were categorized as present or absent for each category

**Area deprivation index (ADI) is a measure of socioeconomic disadvantage and was grouped into quintiles from least to most deprived neighborhoods (based on zip code).

***Frailty was defined using the ICD-10 updated Veterans Affairs Frailty Index (VA-FI) and categorized as non-frail (VA-FI ≤ 0.1), prefrail (>0.1–0.2), mildly frail (>0.2–0.3), moderately frail (>0.3–0.4), and severely frail (>0.5).

****Immunocompromised status was based on immunocompromising conditions in the year prior and immunosuppressive medications in the 90 days prior to the ARI episode based on a slightly modified algorithm that has been previously described. Unlike the previously described algorithm, we used diagnosis codes to identify solid organ or hematopoietic stem cell transplantation and HIV/AIDs versus patient registries.  Consistent with the previously described algorithm, we required one inpatient or two outpatient diagnosis code for an immunocompromising condition (leukemia, lymphoma, congenital immunodeficiencies, asplenia/hyposplenia, HIV/AIDS, and organ transplant) in the year prior and any immunosuppressive medication (alkylating agents, antibiotics, antimetbolites, antimitotics, monoclonal antibodies, other, immune-modulating agents, TNF Alpha antagonist, and steroids) with an outpatient days supply or inpatient administration in the 90 days prior.

****Prior COVID-19 infection was defined as any previous documented SARS-CoV-2 infection or no prior documented infection (yes or no).

*****Virtual visit was only assessed among those with an outpatient visit and defined as a virtual visit or not .

*****ICU admission was only assessed among those with a hospital admission and defined as admission to an ICU or not.

**Supplemental Table 4.** Demographics and clinical characteristics of acute respiratory infection ED/UC visits by case-control status

|  | **Total (n=23,966)** | **SARS-CoV-2 positive**  **(n=4,655)** | **SARS-CoV-2 negative**  **(n=19,311)** | ***P*-value** |
| --- | --- | --- | --- | --- |
| COVID vaccine status*  ≥1 dose of BNT162b2 KP.2 vaccine | 869 (3.6) | 93 (2) | 776 (4.0) | <0.001 |
| ≥1 dose of XBB vaccine | 5,784 (24.1) | 1,216 (26.1) | 4,568 (23.7) | <0.001 |
| ≥1 dose of BA.4/5-adapted bivalent vaccine | 6,890 (28.7) | 1,536 (33.0) | 5,354 (27.7) | <0.001 |
| ≥3 doses of original wild-type mRNA vaccine but no variant-adapted vaccines | 6,342 (26.5) | 1,298 (27.9) | 5,044 (26.1) | 0.014 |
| ≥2 doses of original wild-type mRNA vaccine but no variant-adapted vaccines | 10,892 (45.4) | 2,109 (45.3) | 8,783 (45.5) | 0.829 |
| Unvaccinated | 4,598 (19.2) | 721 (15.5) | 3,877 (20.1) | <0.001 |
| Time since last non-KP.2 adapted vaccine, median days (IQR) | 878 (370-1,102) | 825 (351-1,076) | 889 (374-1,108) | <0.001 |
| Age group  18–64 years | 11,933 (49.8) | 2,039 (43.8) | 9,894 (51.2) | <0.001 |
| 65–74 years | 5,659 (23.6) | 1,097 (23.6) | 4,562 (23.6) |  |
| ≥75 years | 6,374 (26.6) | 1,519 (32.6) | 4,855 (25.1) |  |
| Sex  Male | 20,522 (85.6) | 4,033 (86.6) | 16,489 (85.4) | 0.029 |
| Female | 3,444 (14.4) | 622 (13.4) | 2,822 (14.6) |  |
| Body mass index category  Underweight (<18.5 kg/m^2^) | 184 (0.8) | 28 (0.6) | 156 (0.8) | 0.002 |
| Healthy weight (18.5–24.9 kg/m^2^) | 6,497 (27.1) | 1,266 (27.2) | 5,231 (27.1) |  |
| Overweight (25–29.9 kg/m^2^) | 4,901 (20.4) | 1,040 (22.3) | 3,861 (20.0) |  |
| Obese (≥30 kg/m^2^) | 12,334 (51.5) | 2,314 (49.7) | 10,020 (51.9) |  |
| Missing | 50 (0.2) | 7 (0.2) | 43 (0.2) |  |
| Region  Midwest | 4,836 (20.2) | 976 (21.0) | 3,860 (20.0) | <0.001 |
| Northeast | 3,466 (14.5) | 730 (15.7) | 2,736 (14.2) |  |
| West | 4,593 (19.2) | 1,002 (21.5) | 3,591 (18.6) |  |
| South | 11,071 (46.2) | 1,947 (41.8) | 9,124 (47.2) |  |
| Race  Black or African American | 6,842 (28.5) | 1,190 (25.6) | 5,652 (29.3) | <0.001 |
| White | 14,531 (60.6) | 2,987 (64.2) | 11,544 (59.8) |  |
| Other race | 2,593 (10.8) | 478 (10.3) | 2,115 (11.0) |  |
| Ethnicity  Hispanic or Latino | 2,443 (10.2) | 409 (8.8) | 2,034 (10.5) | <0.001 |
| Not Hispanic or Latino | 21,523 (89.8) | 4,246 (91.2) | 17,277 (89.5) |  |
| Smoking  Current or former | 13,454 (56.1) | 2,540 (54.6) | 10,914 (56.5) | 0.003 |
| Never smoked | 10,055 (42.0) | 2,043 (43.9) | 8,012 (41.5) |  |
| Unknown | 457 (1.9) | 72 (1.5) | 385 (2.0) |  |
| Area deprivation index (ADI)* Quintile  1 (Least Deprived) | 4,638 (19.4) | 839 (18.0) | 3,799 (19.7) | <0.001 |
| 2 | 4,748 (19.8) | 973 (20.9) | 3,775 (19.5) |  |
| 3 | 4,684 (19.5) | 884 (19.0) | 3,800 (19.7) |  |
| 4 | 4,688 (19.6) | 964 (20.7) | 3,724 (19.3) |  |
| 5 (Most Deprived) | 4,447 (18.6) | 897 (19.3) | 3,550 (18.4) |  |
| Missing | 761 (3.2) | 98 (2.1) | 663 (3.4) |  |
| VA Frailty index (VA-FI)**  Non-frail (VA-FI *≤*0.1) | 7,956 (33.2) | 1,489 (32) | 6,467 (33.5) | <0.001 |
| Pre-frail (VA-FI >0.1-0.2) | 6,376 (26.6) | 1,320 (28.4) | 5,056 (26.2) |  |
| Mildly frail (VA-FI >0.2-0.3) | 4,303 (18) | 899 (19.3) | 3,404 (17.6) |  |
| Moderately frail (VA-FI >0.3-0.4) | 2,676 (11.2) | 510 (11.0) | 2,166 (11.2) |  |
| Severely frail (VA-FI ≥0.5) | 2,655 (11.1) | 437 (9.4) | 2,218 (11.5) |  |
| Healthcare exposures, 1 year prior  Hospital admission | 5,130 (21.4) | 771 (16.6) | 4,359 (22.6) | <0.001 |
| Nursing home admission | 521 (2.2) | 76 (1.6) | 445 (2.3) | 0.005 |
| Intensive care unit admission | 1,221 (5.1) | 137 (2.9) | 1,084 (5.6) | <0.001 |
| Emergency department visit | 16,770 (70) | 3,070 (66.0) | 13,700 (70.9) | <0.001 |
| Primary care visit | 22,960 (95.8) | 4,482 (96.3) | 18,478 (95.7) | 0.068 |
| Charlson Comorbidity Index  0 | 8,087 (33.7) | 1,506 (32.4) | 6,581 (34.1) | <0.001 |
| 1 | 4,954 (20.7) | 1,022 (22.0) | 3,932 (20.4) |  |
| 2 | 2,866 (12.0) | 610 (13.1) | 2,256 (11.7) |  |
| 3 | 2,380 (9.9) | 502 (10.8) | 1,878 (9.7) |  |
| ≥4 | 5,679 (23.7) | 1,015 (21.8) | 4,664 (24.2) |  |
| Immunocompromised*** | 9,508 (39.7) | 1,455 (31.3) | 8,053 (41.7) | <0.001 |
| Week of ARI  Sep 05–Sep 07,2023 | 1,178 (4.9) | 456 (9.8) | 722 (3.7) | <0.001 |
| Sep 08–Sep 14,2024 | 2,510 (10.5) | 901 (19.4) | 1,609 (8.3) |  |
| Sep 15–Sep 21,2024 | 2,001 (8.3) | 653 (14.0) | 1,348 (7.0) |  |
| Sep 22–Sep 28,2024 | 1,784 (7.4) | 419 (9.0) | 1,365 (7.1) |  |
| Sep 29–Oct 05,2024 | 1,753 (7.3) | 350 (7.5) | 1,403 (7.3) |  |
| Oct 06–Oct 12,2024 | 1,540 (6.4) | 245 (5.3) | 1,295 (6.7) |  |
| Oct 13–Oct 19,2024 | 1,644 (6.9) | 235 (5.0) | 1,409 (7.3) |  |
| Oct 20–Oct 26,2024 | 1,715 (7.2) | 223 (4.8) | 1,492 (7.7) |  |
| Oct 27–Nov 02,2024 | 1,708 (7.1) | 220 (4.7) | 1,488 (7.7) |  |
| Nov 03–Nov 09,2024 | 1,818 (7.6) | 250 (5.4) | 1,568 (8.1) |  |
| Nov 10–Nov 16,2024 | 1,967 (8.2) | 223 (4.8) | 1,744 (9.0) |  |
| Nov 17–Nov 23,2024 | 2,219 (9.3) | 226 (4.9) | 1,993 (10.3) |  |
| Nov 24–Nov 30,2024 | 2,129 (8.9) | 254 (5.5) | 1,875 (9.7) |  |
| Prior COVID-19 infection**** | 9,337 (39) | 1,644 (35.3) | 7,693 (39.8) | <0.001 |
| Current influenza vaccine | 4,603 (19.2) | 688 (14.8) | 3,915 (20.3) | <0.001 |
| Pneumococcal vaccine in last 5 years | 8,933 (37.3) | 1,734 (37.3) | 7,199 (37.3) | 0.971 |

ARI= acute respiratory infection; ED/UC= emergency department/urgent care; VA= Veterans Affairs

Data are n (%) unless otherwise specified.

All ARI encounters within a 30-day window were considered a single ARI episode. If multiple encounter types occurred during the 30-day window, the highest level of care was used (hospitalization > ED/UC > outpatient).

*The categories under “COVID vaccine status” were categorized as present or absent for each category

**Area deprivation index (ADI) is a measure of socioeconomic disadvantage and was grouped into quintiles from least to most deprived neighborhoods (based on zip code).

***Frailty was defined using the ICD-10 updated Veterans Affairs Frailty Index (VA-FI) and categorized as non-frail (VA-FI ≤ 0.1), prefrail (>0.1–0.2), mildly frail (>0.2–0.3), moderately frail (>0.3–0.4), and severely frail (>0.5).

****Immunocompromised status was based on immunocompromising conditions in the year prior and immunosuppressive medications in the 90 days prior to the ARI episode based on a slightly modified algorithm that has been previously described. Unlike the previously described algorithm, we used diagnosis codes to identify solid organ or hematopoietic stem cell transplantation and HIV/AIDs versus patient registries.  Consistent with the previously described algorithm, we required one inpatient or two outpatient diagnosis code for an immunocompromising condition (leukemia, lymphoma, congenital immunodeficiencies, asplenia/hyposplenia, HIV/AIDS, and organ transplant) in the year prior and any immunosuppressive medication (alkylating agents, antibiotics, antimetbolites, antimitotics, monoclonal antibodies, other, immune-modulating agents, TNF Alpha antagonist, and steroids) with an outpatient days supply or inpatient administration in the 90 days prior.

****Prior COVID-19 infection was defined as any previous documented SARS-CoV-2 infection or no prior documented infection (yes or no).

*****Virtual visit was only assessed among those with an outpatient visit and defined as a virtual visit or not .

*****ICU admission was only assessed among those with a hospital admission and defined as admission to an ICU or not.

**Supplemental Table 5.** Demographics and clinical characteristics of acute respiratory infection outpatient visits by case-control status

|  | **Total (n=11,193)** | **SARS-CoV-2 positive**  **(n=1,677)** | **SARS-CoV-2 negative**  **(n=9,516)** | ***P*-value** |
| --- | --- | --- | --- | --- |
| COVID vaccine status*  ≥1 dose of BNT162b2 KP.2 vaccine | 464 (4.1) | 33 (2.0) | 431 (4.5) | <0.001 |
| ≥1 dose of XBB vaccine | 3,093 (27.6) | 471 (28.1) | 2,622 (27.6) | 0.653 |
| ≥1 dose of BA.4/5-adapted bivalent vaccine | 3,519 (31.4) | 571 (34.0) | 2,948 (31.0) | 0.013 |
| ≥3 doses of original wild-type mRNA vaccine but no variant-adapted vaccines | 2,921 (26.1) | 454 (27.1) | 2,467 (25.9) | 0.324 |
| ≥2 doses of original wild-type mRNA vaccine but no variant-adapted vaccines | 4,845 (43.3) | 725 (43.2) | 4,120 (43.3) | 0.961 |
| Unvaccinated | 2,081 (18.6) | 277 (16.5) | 1,804 (19.0) | 0.018 |
| Time since last non-KP.2 adapted vaccine, median days (IQR) | 785 (348-1,081) | 776 (343-1,070) | 785 (349-1,084) | 0.287 |
| Age group  18–64 years | 4,664 (41.7) | 726 (43.3) | 3,938 (41.4) | 0.278 |
| 65–74 years | 2,927 (26.2) | 417 (24.9) | 2,510 (26.4) |  |
| ≥75 years | 3,602 (32.2) | 534 (31.8) | 3,068 (32.2) |  |
| Sex  Male | 9,708 (86.7) | 1,441 (85.9) | 8,267 (86.9) | 0.292 |
| Female | 1,485 (13.3) | 236 (14.1) | 1,249 (13.1) |  |
| Body mass index category  Underweight (<18.5 kg/m^2^) | 148 (1.3) | 8 (0.5) | 140 (1.5) | <0.001 |
| Healthy weight (18.5–24.9 kg/m^2^) | 3,542 (31.6) | 472 (28.1) | 3,070 (32.3) |  |
| Overweight (25–29.9 kg/m^2^) | 2,304 (20.6) | 388 (23.1) | 1,916 (20.1) |  |
| Obese (≥30 kg/m^2^) | 5,181 (46.3) | 805 (48.0) | 4,376 (46.0) |  |
| Missing | 18 (0.2) | <5 (<0.3) | 14 (0.1) |  |
| Region  Midwest | 2,362 (21.1) | 362 (21.6) | 2,000 (21.0) | 0.233 |
| Northeast | 1,474 (13.2) | 236 (14.1) | 1,238 (13.0) |  |
| West | 2,028 (18.1) | 318 (19.0) | 1,710 (18.0) |  |
| South | 5,329 (47.6) | 761 (45.4) | 4,568 (48.0) |  |
| Race  Black or African American | 2,682 (24.0) | 362 (21.6) | 2,320 (24.4) | 0.032 |
| White | 7,340 (65.6) | 1,124 (67.0) | 6,216 (65.3) |  |
| Other race | 1,171 (10.5) | 191 (11.4) | 980 (10.3) |  |
| Ethnicity  Hispanic or Latino | 956 (8.5) | 159 (9.5) | 797 (8.4) | 0.135 |
| Not Hispanic or Latino | 10,237 (91.5) | 1,518 (90.5) | 8,719 (91.6) |  |
| Smoking  Current or former | 6,714 (60.0) | 962 (57.4) | 5,752 (60.4) | 0.036 |
| Never smoked | 4,192 (37.5) | 675 (40.3) | 3,517 (37.0) |  |
| Unknown | 287 (2.6) | 40 (2.4) | 247 (2.6) |  |
| Area deprivation index (ADI)* Quintile  1 (Least Deprived) | 2,219 (19.8) | 324 (19.3) | 1,895 (19.9) | 0.523 |
| 2 | 2,219 (19.8) | 336 (20.0) | 1,883 (19.8) |  |
| 3 | 2,153 (19.2) | 340 (20.3) | 1,813 (19.1) |  |
| 4 | 2,139 (19.1) | 326 (19.4) | 1,813 (19.1) |  |
| 5 (Most Deprived) | 2,261 (20.2) | 316 (18.8) | 1,945 (20.4) |  |
| Missing | 202 (1.8) | 35 (2.1) | 167 (1.8) |  |
| VA Frailty index (VA-FI)**  Non-frail (VA-FI *≤*0.1) | 2,571 (23.0) | 489 (29.2) | 2,082 (21.9) | <0.001 |
| Pre-frail (VA-FI >0.1-0.2) | 2,493 (22.3) | 475 (28.3) | 2,018 (21.2) |  |
| Mildly frail (VA-FI >0.2-0.3) | 2,174 (19.4) | 344 (20.5) | 1,830 (19.2) |  |
| Moderately frail (VA-FI >0.3-0.4) | 1,651 (14.8) | 191 (11.4) | 1,460 (15.3) |  |
| Severely frail (VA-FI ≥0.5) | 2,304 (20.6) | 178 (10.6) | 2,126 (22.3) |  |
| Healthcare exposures, 1 year prior  Hospital admission | 4,685 (41.9) | 364 (21.7) | 4,321 (45.4) | <0.001 |
| Nursing home admission | 911 (8.1) | 166 (9.9) | 745 (7.8) | 0.004 |
| Intensive care unit admission | 1,783 (15.9) | 101 (6.0) | 1,682 (17.7) | <0.001 |
| Emergency department visit | 7,106 (63.5) | 799 (47.6) | 6,307 (66.3) | <0.001 |
| Primary care visit | 10,635 (95.0) | 1,597 (95.2) | 9,038 (95.0) | 0.661 |
| Charlson Comorbidity Index  0 | 2,732 (24.4) | 562 (33.5) | 2,170 (22.8) | <0.001 |
| 1 | 1,890 (16.9) | 328 (19.6) | 1,562 (16.4) |  |
| 2 | 1,367 (12.2) | 208 (12.4) | 1,159 (12.2) |  |
| 3 | 1,193 (10.7) | 195 (11.6) | 998 (10.5) |  |
| ≥4 | 4,011 (35.8) | 384 (22.9) | 3,627 (38.1) |  |
| Immunocompromised*** | 4,058 (36.3) | 433 (25.8) | 3,625 (38.1) | <0.001 |
| Week of ARI  Sep 05–Sep 07,2023 | 1,213 (10.8) | 274 (16.3) | 939 (9.9) | <0.001 |
| Sep 08–Sep 14,2024 | 1,469 (13.1) | 370 (22.1) | 1,099 (11.5) |  |
| Sep 15–Sep 21,2024 | 989 (8.8) | 207 (12.3) | 782 (8.2) |  |
| Sep 22–Sep 28,2024 | 828 (7.4) | 133 (7.9) | 695 (7.3) |  |
| Sep 29–Oct 05,2024 | 807 (7.2) | 121 (7.2) | 686 (7.2) |  |
| Oct 06–Oct 12,2024 | 700 (6.3) | 93 (5.5) | 607 (6.4) |  |
| Oct 13–Oct 19,2024 | 604 (5.4) | 61 (3.6) | 543 (5.7) |  |
| Oct 20–Oct 26,2024 | 742 (6.6) | 88 (5.2) | 654 (6.9) |  |
| Oct 27–Nov 02,2024 | 782 (7.0) | 73 (4.4) | 709 (7.5) |  |
| Nov 03–Nov 09,2024 | 794 (7.1) | 77 (4.6) | 717 (7.5) |  |
| Nov 10–Nov 16,2024 | 662 (5.9) | 56 (3.3) | 606 (6.4) |  |
| Nov 17–Nov 23,2024 | 874 (7.8) | 75 (4.5) | 799 (8.4) |  |
| Nov 24–Nov 30,2024 | 729 (6.5) | 49 (2.9) | 680 (7.1) |  |
| Prior COVID-19 infection**** | 3,754 (33.5) | 529 (31.5) | 3,225 (33.9) | 0.061 |
| Virtual visit (outpatient only)***** | 979 (8.7) | 401 (23.9) | 578 (6.1) | <0.001 |
| Current influenza vaccine | 2,148 (19.2) | 237 (14.1) | 1,911 (20.1) | <0.001 |
| Pneumococcal vaccine in last 5 years | 4,447 (39.7) | 646 (38.5) | 3,801 (39.9) | 0.272 |

ARI= acute respiratory infection; ED/UC= emergency department/urgent care; VA= Veterans Affairs

Data are n (%) unless otherwise specified.

All ARI encounters within a 30-day window were considered a single ARI episode. If multiple encounter types occurred during the 30-day window, the highest level of care was used (hospitalization > ED/UC > outpatient).

*The categories under “COVID vaccine status” were categorized as present or absent for each category

**Area deprivation index (ADI) is a measure of socioeconomic disadvantage and was grouped into quintiles from least to most deprived neighborhoods (based on zip code).

***Frailty was defined using the ICD-10 updated Veterans Affairs Frailty Index (VA-FI) and categorized as non-frail (VA-FI ≤ 0.1), prefrail (>0.1–0.2), mildly frail (>0.2–0.3), moderately frail (>0.3–0.4), and severely frail (>0.5).

****Immunocompromised status was based on immunocompromising conditions in the year prior and immunosuppressive medications in the 90 days prior to the ARI episode based on a slightly modified algorithm that has been previously described. Unlike the previously described algorithm, we used diagnosis codes to identify solid organ or hematopoietic stem cell transplantation and HIV/AIDs versus patient registries.  Consistent with the previously described algorithm, we required one inpatient or two outpatient diagnosis code for an immunocompromising condition (leukemia, lymphoma, congenital immunodeficiencies, asplenia/hyposplenia, HIV/AIDS, and organ transplant) in the year prior and any immunosuppressive medication (alkylating agents, antibiotics, antimetbolites, antimitotics, monoclonal antibodies, other, immune-modulating agents, TNF Alpha antagonist, and steroids) with an outpatient days supply or inpatient administration in the 90 days prior.

****Prior COVID-19 infection was defined as any previous documented SARS-CoV-2 infection or no prior documented infection (yes or no).

*****Virtual visit was only assessed among those with an outpatient visit and defined as a virtual visit or not .

*****ICU admission was only assessed among those with a hospital admission and defined as admission to an ICU or not.

**Supplemental Table 6.** Demographics and clinical characteristics of acute respiratory infection encounters by BNT162b2 KP.2 vaccination status

| **Variable** | **Total (n=44,598)** | **Received BNT162b2 KP.2 vaccine (n=1,666)** | **No KP.2 vaccine of any kind**  **(n=42,932)** | ***P*-value** |
| --- | --- | --- | --- | --- |
| COVID vaccine status  ≥1 dose of BNT162b2 XBB vaccine | 11,786 (26.4) | 1,277 (76.7) | 10,509 (24.5) | <0.001 |
| ≥1 dose of BA.4/5-adapted bivalent vaccine | 13,730 (30.8) | 1,258 (75.5) | 12,472 (29.1) | <0.001 |
| ≥3 doses of original wild-type mRNA vaccine but no variant-adapted vaccines | 11,916 (26.7) | 303 (18.2) | 11,613 (27.0) | <0.001 |
| ≥2 doses of original wild-type mRNA vaccine but no variant-adapted vaccines | 19,819 (44.4) | 364 (21.8) | 19,455 (45.3) | <0.001 |
| Unvaccinated | 8,162 (18.3) | 14 (0.8) | 8,148 (19) | <0.001 |
| Time since last non-KP.2 adapted vaccine, median days (IQR) | 815 (354-1,088) | 376 (327-410) | 856 (357-1,095) | <0.001 |
| Age group  18–64 years | 18,447 (41.4) | 328 (19.7) | 18,119 (42.2) | <0.001 |
| 65–74 years | 11,583 (26) | 529 (31.8) | 11,054 (25.7) |  |
| ≥75 years | 14,568 (32.7) | 809 (48.6) | 13,759 (32.0) |  |
| Sex  Male | 39,127 (87.7) | 1,497 (89.9) | 37,630 (87.7) | 0.007 |
| Female | 5,471 (12.3) | 169 (10.1) | 5,302 (12.3) |  |
| Body mass index category  Underweight (<18.5 kg/m^2^) | 657 (1.5) | 17 (1.0) | 640 (1.5) | 0.022 |
| Healthy weight (18.5–24.9 kg/m^2^) | 14,107 (31.6) | 580 (34.8) | 13,527 (31.5) |  |
| Overweight (25–29.9 kg/m^2^) | 8,883 (19.9) | 336 (20.2) | 8,547 (19.9) |  |
| Obese (≥30 kg/m^2^) | 20,877 (46.8) | 731 (43.9) | 20,146 (46.9) |  |
| Missing | 74 (0.2) | <5 (<0.3) | 72 (0.2) |  |
| Region  Midwest | 9,005 (20.2) | 434 (26.1) | 8,571 (20.0) | <0.001 |
| Northeast | 6,208 (13.9) | 308 (18.5) | 5,900 (13.7) |  |
| West | 8,773 (19.7) | 353 (21.2) | 8,420 (19.6) |  |
| South | 20,612 (46.2) | 571 (34.3) | 20,041 (46.7) |  |
| Race  Black or African American | 11,760 (26.4) | 514 (30.9) | 11,246 (26.2) | <0.001 |
| White | 28,269 (63.4) | 1,007 (60.4) | 27,262 (63.5) |  |
| Other race | 4,569 (10.2) | 145 (8.7) | 4,424 (10.3) |  |
| Ethnicity  Hispanic or Latino | 4,120 (9.2) | 92 (5.5) | 4,028 (9.4) | <0.001 |
| Not Hispanic or Latino | 40,478 (90.8) | 1,574 (94.5) | 38,904 (90.6) |  |
| Smoking  Current or former | 26,594 (59.6) | 1,025 (61.5) | 25,569 (59.6) | 0.039 |
| Never smoked | 17,007 (38.1) | 617 (37.0) | 16,390 (38.2) |  |
| Unknown | 997 (2.2) | 24 (1.4) | 973 (2.3) |  |
| Area deprivation index (ADI) Quintile  1 (Least Deprived) | 8,671 (19.4) | 464 (27.9) | 8,207 (19.1) | <0.001 |
| 2 | 8,666 (19.4) | 354 (21.2) | 8,312 (19.4) |  |
| 3 | 8,668 (19.4) | 284 (17.0) | 8,384 (19.5) |  |
| 4 | 8,683 (19.5) | 245 (14.7) | 8,438 (19.7) |  |
| 5 (Most Deprived) | 8,668 (19.4) | 307 (18.4) | 8,361 (19.5) |  |
| Missing | 1,242 (2.8) | 12 (0.7) | 1,230 (2.9) |  |
| VA Frailty index (VA-FI)*  Non-frail (VA-FI *≤*0.1) | 11,359 (25.5) | 186 (11.2) | 11,173 (26.0) | <0.001 |
| Pre-frail (VA-FI >0.1-0.2) | 10,353 (23.2) | 371 (22.3) | 9,982 (23.3) |  |
| Mildly frail (VA-FI >0.2-0.3) | 8,452 (19.0) | 375 (22.5) | 8,077 (18.8) |  |
| Moderately frail (VA-FI >0.3-0.4) | 6,305 (14.1) | 303 (18.2) | 6,002 (14.0) |  |
| Severely frail (VA-FI ≥0.5) | 8,129 (18.2) | 431 (25.9) | 7,698 (17.9) |  |
| Healthcare exposures, 1 year prior  Hospital admission | 15,243 (34.2) | 682 (40.9) | 14,561 (33.9) | <0.001 |
| Nursing home admission | 2,027 (4.5) | 80 (4.8) | 1,947 (4.5) | 0.608 |
| Intensive care unit admission | 4,877 (10.9) | 210 (12.6) | 4,667 (10.9) | 0.026 |
| Emergency department visit | 31,389 (70.4) | 1,243 (74.6) | 30,146 (70.2) | <0.001 |
| Primary care visit | 42,629 (95.6) | 1,627 (97.7) | 41,002 (95.5) | <0.001 |
| Charlson Comorbidity Index  0 | 11,627 (26.1) | 228 (13.7) | 11,399 (26.6) | <0.001 |
| 1 | 8,026 (18.0) | 276 (16.6) | 7,750 (18.1) |  |
| 2 | 5,370 (12.0) | 213 (12.8) | 5,157 (12.0) |  |
| 3 | 4,778 (10.7) | 226 (13.6) | 4,552 (10.6) |  |
| ≥4 | 14,797 (33.2) | 723 (43.4) | 14,074 (32.8) |  |
| Immunocompromised** | 18,223 (40.9) | 743 (44.6) | 17,480 (40.7) | 0.002 |
| Medical History***  Acute cerebrovascular disease | 2,673 (6.0) | 120 (7.2) | 2,553 (5.9) | 0.034 |
| Acute myocardial infarction | 2,035 (4.6) | 102 (6.1) | 1,933 (4.5) | 0.002 |
| Alcohol and substance related disorders | 12,323 (27.6) | 401 (24.1) | 11,922 (27.8) | <0.001 |
| Any cancer or malignancy | 19,271 (43.2) | 910 (54.6) | 18,361 (42.8) | <0.001 |
| Aortic and peripheral arterial embolism or thrombosis | 246 (0.6) | 13 (0.8) | 233 (0.5) | 0.199 |
| Asthma | 4,590 (10.3) | 212 (12.7) | 4,378 (10.2) | <0.001 |
| Benign prostatic hyperplasia | 11,427 (25.6) | 584 (35.1) | 10,843 (25.3) | <0.001 |
| Cardiac dysrhythmias | 13,229 (29.7) | 635 (38.1) | 12,594 (29.3) | <0.001 |
| Chronic kidney disease | 6,642 (14.9) | 332 (19.9) | 6,310 (14.7) | <0.001 |
| Chronic obstructive pulmonary disease and bronchiectasis | 12,986 (29.1) | 623 (37.4) | 12,363 (28.8) | <0.001 |
| Congestive heart failure | 9,040 (20.3) | 431 (25.9) | 8,609 (20.1) | <0.001 |
| Coronary atherosclerosis and other heart disease | 11,881 (26.6) | 591 (35.5) | 11,290 (26.3) | <0.001 |
| Delirium, dementia, and other cognitive disorders | 5,170 (11.6) | 251 (15.1) | 4,919 (11.5) | <0.001 |
| Diabetes with or without chronic complications | 20,884 (46.8) | 938 (56.3) | 19,946 (46.5) | <0.001 |
| Epilepsy | 1,520 (3.4) | 86 (5.2) | 1,434 (3.3) | <0.001 |
| Human immunodeficiency virus (HIV) infection | 593 (1.3) | 45 (2.7) | 548 (1.3) | <0.001 |
| Hypertension | 28,801 (64.6) | 1,257 (75.5) | 27,544 (64.2) | <0.001 |
| Influenza | 1,245 (2.8) | 30 (1.8) | 1,215 (2.8) | 0.012 |
| Liver diseases | 5,123 (11.5) | 186 (11.2) | 4,937 (11.5) | 0.674 |
| Mental health conditions | 23,855 (53.5) | 875 (52.5) | 22,980 (53.5) | 0.419 |
| Osteoarthritis | 10,803 (24.2) | 514 (30.9) | 10,289 (24.0) | <0.001 |
| Peripheral and visceral atherosclerosis | 4,744 (10.6) | 242 (14.5) | 4,502 (10.5) | <0.001 |
| Pneumonia | 5,829 (13.1) | 253 (15.2) | 5,576 (13.0) | 0.009 |
| Pulmonary heart disease | 4,126 (9.3) | 188 (11.3) | 3,938 (9.2) | 0.004 |
| Rheumatoid arthritis | 1,063 (2.4) | 46 (2.8) | 1,017 (2.4) | 0.303 |
| Septicemia | 3,005 (6.7) | 106 (6.4) | 2,899 (6.8) | 0.533 |
| Thyroid disorder | 6,325 (14.2) | 282 (16.9) | 6,043 (14.1) | 0.001 |
| Tuberculosis | 126 (0.3) | 8 (0.5) | 118 (0.3) | 0.121 |
| Week of ARI  Sep 05–Sep 07,2023 | 2,965 (6.6) | <5 (<0.3) | 2,965 (6.9) | <0.001 |
| Sep 08–Sep 14,2024 | 5,015 (11.2) | <5 (<0.3) | 5,014 (11.7) |  |
| Sep 15–Sep 21,2024 | 3,971 (8.9) | <5 (<0.3) | 3,967 (9.2) |  |
| Sep 22–Sep 28,2024 | 3,498 (7.8) | 21 (1.3) | 3,477 (8.1) |  |
| Sep 29–Oct 05,2024 | 3,398 (7.6) | 48 (2.9) | 3,350 (7.8) |  |
| Oct 06–Oct 12,2024 | 2,963 (6.6) | 55 (3.3) | 2,908 (6.8) |  |
| Oct 13–Oct 19,2024 | 2,997 (6.7) | 119 (7.1) | 2,878 (6.7) |  |
| Oct 20–Oct 26,2024 | 3,215 (7.2) | 158 (9.5) | 3,057 (7.1) |  |
| Oct 27–Nov 02,2024 | 3,207 (7.2) | 202 (12.1) | 3,005 (7) |  |
| Nov 03–Nov 09,2024 | 3,333 (7.5) | 233 (14) | 3,100 (7.2) |  |
| Nov 10–Nov 16,2024 | 3,421 (7.7) | 267 (16) | 3,154 (7.3) |  |
| Nov 17–Nov 23,2024 | 3,658 (8.2) | 312 (18.7) | 3,346 (7.8) |  |
| Nov 24–Nov 30,2024 | 2,957 (6.6) | 246 (14.8) | 2,711 (6.3) |  |
| Prior COVID-19 infection | 16,270 (36.5) | 663 (39.8) | 15,607 (36.4) | 0.004 |
| Virtual visit (outpatient only) | 979 (8.7) | 43 (9.3) | 936 (8.7) | 0.685 |
| ICU admission (hospitalized only) | 2,132 (22.6) | 60 (18.0) | 2,072 (22.8) | 0.042 |
| Current influenza vaccine | 8,731 (19.6) | 1,554 (93.3) | 7,177 (16.7) | <0.001 |
| Pneumococcal vaccine in last 5 years | 17,174 (38.5) | 879 (52.8) | 16,295 (38) | <0.001 |

ARI= acute respiratory infection; ED/UC= emergency department/urgent care; VA= Veterans Affairs

Data are n (%) unless otherwise specified. Chi-square or Fisher’s Exact tests were used to compare differences in proportions between the groups. For continuous variables, comparisons were performed using the Wilcoxon Rank Sum test or a Student’s t-test, depending on the distribution of the data for the given variable.

All ARI encounters within a 30-day window were considered a single ARI episode. If multiple encounter types occurred during the 30-day window, the highest level of care was used (hospitalization > ED/UC > outpatient).

*VA Frailty index was categorized as non-frail (VA-FI ≤ 0.1), prefrail (>0.1–0.2), mildly frail (>0.2–0.3), moderately frail (>0.3–0.4), and severely frail (>0.5).

**Immunocompromised status was based on immunocompromising conditions in the year prior and immunosuppressive medications in the 90 days prior to the ARI episode based on a slightly modified algorithm that has been previously described. (Tartof SY, et al. Lancet Reg Health Am. 2022:9:100198.) Unlike the previously described algorithm, we used diagnosis codes to identify solid organ or hematopoietic stem cell transplantation and HIV/AIDs versus patient registries.  Consistent with the previously described algorithm, we required one inpatient or two outpatient diagnosis code for an immunocompromising condition (leukemia, lymphoma, congenital immunodeficiencies, asplenia/hyposplenia, HIV/AIDS, and organ transplant) in the year prior and any immunosuppressive medication (alkylating agents, antibiotics, antimetbolites, antimitotics, monoclonal antibodies, other, immune-modulating agents, TNF Alpha antagonist, and steroids) with an outpatient days supply or inpatient administration in the 90 days prior.

***Medical history included underlying conditions and diagnoses in the year prior to the ARI episode, identified using international classification of diseases (ICD)-10 codes.

**Supplemental Table 7.** Adjusted effectiveness of the BNT162b2 KP.2 vaccine by COVID-19 outcomes among those ≥65 years of age

| **Outcome** | **Age ≥65 years**  **(n = 26,151)** | |
| --- | --- | --- |
|  | **VE (95% CI)** | **Median time since KP.2 vaccine, days (IQR)** |
| Hospitalization | 75 (49–88) | 30 (22-43) |
| ED/UC visit | 56 (44–66) | 36 (25-49) |
| Outpatient visit | 58 (36–73) | 31 (20-46) |

ED/UC = emergency department/urgent care; IQR = interquartile range; KP.2 = BNT162b2 KP.2 adapted vaccine; VE= vaccine effectiveness

Compared the odds of receiving the 2024/2025 BNT162b2 KP.2 strain-adapted COVID-19 vaccine between SARS-CoV-2 positive cases and SARS-CoV-2 negative controls. Adjusted for sex (male or female), race (Black, White, or other race), ethnicity (Hispanic or non-Hispanic), body mass index (BMI) categories (underweight, healthy weight, overweight, obese, missing), Charlson Comorbidity Index (0, 1, 2, 3, ≥4), receipt of pneumococcal vaccine in the past 5 years (yes or no), hospital admission, nursing home admission, ED/UC visit, primary care visit; 0 or ≥1 for each), prior documented SARS-CoV-2 infection (yes or no), smoking status (current/former smoker or never smoker/unknown), immunocompromised (yes or no), and Census region (Northeast, Midwest, South, or West).
